# A tool to evaluate the impact of lived experience involvement in research: the Brain and Genomics Hub *Impact Log* protocol

**DOI:** 10.64898/2026.03.04.26347596

**Authors:** Tania Gergel, Talen Wright, Lavenda Geshica, Emily Vicary, Jaycee Kennett, Oli Delgaram-Nejad, Carina Edwards, Hashwin V.S. Ganesh, Thomas Kabir, Chloe L. Harrison, James Heard, Georgia Dash, Catherine Bresner, Ian Jones, Jeremy Hall, Ann John, Neil A. Harrison, James T.R Walters, Sophie E. Legge

## Abstract

**Background:** Despite widespread recognition of the value of lived experience involvement (LEI) in healthcare research and increased LE involvement activity, we lack established implementation methods and instruments for reporting and evaluating impact. We present a protocol for an innovative LE-led *Impact Log* tool and LEI framework, which may help to address some fundamental barriers to LEI. The *Impact Log* will be implemented within a five-year multidisciplinary transdiagnostic research project on severe mental illness and is also designed for wider adaptation and use. We include a short literature review on LEI impact, to provide in-depth background.

**Methods:** The *Impact Log* framework is designed to integrate inclusive impactful LEI throughout all research stages, and to use a short accessible form to record and evaluate impact across three domains: design and delivery; interpersonal and environmental aspects; systems and processes. *Impact Log* design and implementation is led by LE study leads and specialist LE advisory panel, with combined research experience and LE, fully integrated within the wider research team. All Hub research participants will be offered accessible opportunities for remunerated lived experience input, and there will be outreach to ensure diverse representation, aided by the Hub’s charity partners. Data collection and analysis will be LE led and will include iterative analysis to inform continuing development. Diverse formal and informal dissemination throughout the project will maximise wider stakeholder engagement.

**Conclusion:** The potential value of this research is to implement a novel tool and framework for facilitating, recording and evaluating LEI in complex mental health research, which can be adapted for wider use. Strengths in design are LE leadership and cross-cutting LE research integration, incorporation of multiple domains, and a focus on facilitating diversity and inclusion within LEI. Potential limitations for this project and wider adaptation may include limited resources, risk of bias and health challenges.

**Plain Language Summary:** *We have provided a brief lay summary to help people without a research background understand our project.:* This article explains our plan to develop and test a new way of understanding how research changes when people with personal experience of a mental health condition are part of the research-team. We are mental health researchers, including many with direct experience of bipolar and psychosis, working alongside other researchers, clinicians and charities. Our research project aims to better understand what is happening in the brain, body, lives and experiences of people with bipolar and psychosis. Many people believe research is better when it includes the views of people with direct experience of the health condition being studied. This is called “lived experience”. We have developed a structured approach to ensure that people with lived experience are meaningfully involved in our research. We have also created a simple tool, called the *Impact Log*, to record when lived experience members contribute and help us understand how their involvement influences the research.

*Public and patient / lived experience involvement:* Initial development of the *Impact Log* and LEI framework was by LE researchers and the further development and adaptations described below have been LE-led and co-produced by researchers with LE of bipolar and psychosis. The manuscript preparation has been LE-led and co-produced. This co-production strategy integrates researchers with and without direct LE within an equitable research environment recognising all forms of expertise. We therefore hope to provide an example demonstrating the potential of fully co-produced research, led by researchers with LE, despite the health challenges that we face. Full descriptions of LE involvement in this work can be found throughout this paper and, in fuller detail, in the author contributions and details sections and in the GRIPP2 form in Section 4 of the Extended Materials. For a table showing LE and other relevant experience and expertise amongst the authorial team please see “Table 1: Author disclosed information on mental health LE and professional experience” in Author Contributions below. This article is framed within the conventional language and structures of medical research, in order to present a thorough and rigorous explanation of this protocol. We do recognise that this could present a barrier to adaptation of the tools presented below by people with LE who do not have research backgrounds. However, as detailed below, we will be working to provide accessible versions of this work for dissemination amongst non-specialist audiences. The *Impact Log* itself has been designed to maximise accessibility for users.

## 1. Introduction

The value of Lived Experience Involvement (LEI), sometimes known as “service user”, “consumer”, “public and patient” involvement or “participatory” methods, is increasingly recognised and incorporated into design and delivery of health research and service development. The term “co-production” generally refers to transformative and equitable inclusion of people with LE of relevant health condition/s as collaborative partners, sharing leadership and decision-making.(1) Caregiver perspectives, sometimes known as “indirect” LE, are also recognised as valuable, although concerns exist that this indirect viewpoint might supplant direct LE voices, particularly in mental health contexts where substitute decision making and structural disempowerment are common.(2–4)

LEI, initially viewed as an alternative approach, challenging dominant methodological and epistemological paradigms, is progressively becoming absorbed within mainstream medical research, with mental health often leading both structural adaptation and scale of inclusion. Clear evidence of systemic change and increasing LEI exists(2, 5, 6), with LEI increasingly identified as a research requirement by funders, policy-makers, and publishers (3, 7–12).

Initially, LEI was motivated by ethical and disability-rights drive towards inclusion and representation in medical and policy decisions, exemplified by the “nothing about us without us” principle.(11, 13) More recently, the epistemological value of LEI is emphasised, understanding “expertise by experience” as a distinct form of knowledge, inaccessible to those without direct personal LE(4, 5, 9, 14). This is accompanied by recognition of LEI’s transformative potential for systemic and relational change, including increased empowerment and structural inclusion of people with LE and attitudinal shifts, such as stigma reduction amongst researchers, clinicians, and the wider community.(2)

While this increased emphasis on LEI is welcomed, it remains a relatively new branch of research, with significant gaps to address relating to methodology, systems, and culture to facilitate true integration of complex and impactful LEI.(15, 16) A key challenge is addressing a need for standardised and replicable methods, while ensuring that the authenticity and diversity of LE voices is retained – how can we create frameworks reflecting established academic models, while simultaneously retaining flexibility and openness for articulating voices and perspectives from outside these established models? (11, 14, 17–22)

A major challenge is preventing tokenistic LEI.(3, 7, 9, 10, 16, 23–26) This requires recognition of distinct LE roles, skills, and contributions, alongside structural adaptation creating opportunities for LE leadership, decision-making, adequate resources, and cross-cutting involvement.(5, 6, 8, 23, 24, 26–30) Greater inclusivity and representativeness is needed, particularly from groups underrepresented both demographically and relating to particular mental health conditions and severity.(5, 6, 8, 14, 16, 18, 19, 21–23, 31) We need, for example, to involve more people living with severe conditions including bipolar and psychosis. We must also ensure that LE involvement opportunities exist within biomedical and multidisciplinary research domains, not only within psychosocial and qualitative research.(32, 33)

Systemic integration of LEI requires development of robust and reproducible methods for recording and evaluating scope and impact, while also acknowledging that LEI remains a developing field, incorporating diverse approaches and perspectives. Methods for recording and evaluation must be designed to capture breadth and complexity of impact and to enhance understanding, while also being adaptable for diverse study types and environments.(34, 35) Currently, to our knowledge, there is some guidance, but no widely used tool for recording and evaluating the impact of LEI which meets these criteria.(6, 11, 21, 34, 36–39)

The primary aim of this protocol paper is to present a novel co-produced tool, the *Impact Log*, for evaluating the impact of LEI. This is an exploratory project, implementing a LEI impact evaluation tool and laying down a framework for future development and adaptation. The research setting is a complex multidisciplinary mental health research project, the Brain and Genomics Hub (BG Hub) of the UKRI Mental Health Platform. To give an in-depth overview of core contributory ideas in developing the *Impact Log* and its LEI framework, we now include a brief narrative review defining, evaluating, and increasing the impact of LEI, to provide more detail on the background context for this protocol.

### 1.1 : A short literature review on defining, evaluating, and increasing the impact of LEI

We present “impact” here as positive change on people, systems, or process. Previous reviews have noted that LEI impact is often restricted to early research stages and lacks demographic and health diversity. Also noted is the need for increased reporting, evaluation, and consistency, to capture different contexts and domains of impact, including interpersonal.(5, 6, 24, 25, 28, 40–44). This review, conducted by the BG Hub specialist LEAP (Lived Experience Advisory Panel), includes expert review; non-systematic literature search of peer-reviewed journal articles and grey literature; additional reference searches (for more details on search and screening strategy; data extraction; narrative synthesis see Supplementary Materials 5.1/2). We used an iterative deductive methodology, combining guidelines for narrative synthesis in systematic reviews and thematic analysis,(45–47) to identify themes and subthemes relating to defining, evaluating, and improving LEI impact within extracted data.

We identified 91 peer-reviewed journal articles including reviews, studies, and methodological papers, and five grey literature items, including two sets of guidelines, two comment pieces, and a workshop report. Most items originated in Europe, North America, and Australasia, particularly the United Kingdom, Australia, and Canada. However, we also included individual studies from Brazil, Ethiopia, Nepal, and Israel. Our results are summarised in Table 2, with a full table of themes, subthemes, articles, and references for included articles included in Extended Materials 5.3/4.

**Table 1:** Author disclosed information on mental health LE and professional experience.

| Author | LE of bipolar and/or psychosis | LE of other severe mental illness | LE of supporting family/friend with severe mental illness | Mental health research experience? | Mental health clinical experience? | Experience of MH charity sector work? |
| --- | --- | --- | --- | --- | --- | --- |
| B | Yes | No | Yes | Yes | No | Yes |
| B | Yes | Yes | No | Yes | Yes | Yes |
| B | No | No | No | Yes | No | No |
| B | Yes | No | No | Yes | No | Yes |
| B | No | No | No | Yes | No | Yes |
| B | Yes | Yes | No | Yes | No | No |
| B | No | No | No | Yes | No | Yes |
| B | Yes | No | Yes | Yes | No | No |
| B | No | No | Yes | Yes | Yes | Yes |
| B | No | No | Yes | Yes | Yes | No |
| B | No | No | Yes | Yes | No | No |
| B | No | No | No | Yes | No | Yes |
| B | No | No | No | Yes | No | Yes |
| B | Yes | Yes | No | Yes | No | Yes |
| B | Yes | Yes | Yes | Yes | No | Yes |
| B | Yes | No | Yes | Yes | No | Yes |
| B | No | No | Yes | Yes | Yes | No |
| B | No | No | No | Yes | Yes | No |
| B | No | No | No | Yes | Yes | Yes |

**Table 2:** Summary of themes and subthemes for defining, evaluating, and increasing LEI impact. Abbreviations: LEI = Lived Experience Involvement LE = Lived Experience

| Theme | Summary of themes/subtheme |
| --- | --- |
| **How is impact defined and understood?** |  |
| Theme 1: Change to research / service processes and outcomes | Impact was understood as an increase in quality, relevance, and impact of research and interventions; systemic changes to related institutional structures, processes, and policies, manifesting in areas such as leadership and resources; and changes occurring throughout all stages in the design and delivery of research or services. |
| Theme 2: Interpersonal and cultural change | This focused on manifestation of impact through changed attitudes, such as stigma reduction and increased acceptance amongst researchers and clinicians towards mental illness and LEI. |
| **Methods of impact evaluation** |  |
| Theme 1: Methodologies for assessing impact | This was the most dominant theme, within which the most dominant subtheme was “qualitative reflections”. This emphasised the need to gather primarily reflective and qualitative data, often without clarity about whether these reflections should come from LE or non-LE researchers. |
| Theme 2: Process indicators | This suggested that impact could be measured through assessing tangible systemic changes. |
| Theme 3: The need for alternative ways of understanding impact | This referred to measuring impact from outside conventional psychiatric evaluative frameworks or through examining evidence of critique and change to conventional models and reduced stigma. |
| Theme 4: Assessing impact of co-production on outcomes and dissemination | This suggested that impact could be measured through assessing tangible changes in quality of outcomes and dissemination. |

| **How to increase impact: (multiple diverse subthemes)** |  |
| --- | --- |
| Theme 1: Changing systems and environment | A common suggestion was environmental adaptation to facilitate support, power-sharing, building trust and flexibility. This included systemic changes to research structures, such as LE leadership, co-authorship, establishing academic LE posts, changes to language, offering payments for involvement work, and sustainability. Other subthemes were the need to provide training for both LE and non-LE researchers and to report and evaluate LEI, to increase transparency, adoption, and recognition. |
| Theme 2: Recognising the value and diversity of lived experience involvement | The most common subtheme was the need for true integration of LE researchers, moving away from tokenism and stigma, with clarity, transparency, security, and clear communications surrounding roles and expectations. Another recurrent subtheme was the need for LEI to extend through all stages of research or service design and delivery. Two other subthemes related to making LEI more representative, through including diverse demographics and health conditions and through creating safe and valued spaces for LE contributors to maintain authenticity and facilitate critique. |

Our review highlights wide recognition of the importance of facilitation, reporting, and evaluating impact of LEI impact and that this will require major systemic change including: feasible and equitable LE employment, through broader support and flexibility, and specifics such as reimbursement, leadership, and authorship. Conventional research practices are viewed as insufficient to facilitate truly representative, diverse, and authentic LEI involvement and allow its full potential impact. We must recognise the importance of interpersonal and cultural shifts resulting from LEI, including changes in researcher attitudes and models, power structures, and inclusion. A key related theme was the importance of integrating qualitative experiential reflections, with some suggestions that impact should be evaluated through explicitly assessing evidence of critique and conceptual change.

In conclusion, it appears that there are three well-established core domains relevant to all aspects of impact, which should all be captured in the design of LEI strategies and impact evaluation tools to reflect the true breadth and complexity of impact. These domains are: change in design, delivery, and outcomes; systemic change in processes and environment; interpersonal and cultural change within the research community.(42) However, our review also highlighted the well-recognised challenge and conceptual divide. Despite recognised needs for standardised and replicable methods for implementing, recording, and evaluating LEI, close adherence to conventional research models risks compromising LEI’s authenticity and value. There may not be a perfect solution. Nevertheless, work on LEI must be cognizant of this challenge and transparent about any resulting limitations.

### 2. The Brain and Genomics Hub LEI *Impact Log* protocol

#### 2.1 Methods /Design

##### 2.1.1 Aims

The BG Hub *Impact Log* aims to implement a novel tool to collect data related to impact of LEI on research design, delivery, processes, interpersonal and environmental aspects, adaptable for diverse research contexts. *Impact Log* development is LE-led and co-produced. We hope that *Impact Log* data analysis will help expand understanding of the nature and evaluation of LEI “impact” in complex mental health research. The *Impact Log* tool will be implemented in the BG Hub, a 5-year multidisciplinary and transdiagnostic study of bipolar disorder, schizophrenia, and schizoaffective disorder. It is embedded in the Hub’s wider LEI framework, with core aims of increasing LEI diversity and inclusion and facilitating impactful LEI throughout all research stages. We hope that the *Impact Log* framework will facilitate increased LEI in complex and multidisciplinary mental health research, including biomedically-orientated components and research on severe mental illness.

##### 2.1.2 Study setting: The Brain and Genomics Hub

The BG Hub is a transdiagnostic co-produced investigation of clinical, social, developmental, biomedical (neuroscientific/genomic), and LE factors relating to severe mental illness, aiming to transform understanding, diagnosis and management. One of six new Mental Health Platform research Hubs, established in 2024 by the MRC and UKRI, it unites an interdisciplinary community of researchers and clinicians from South Wales and the Southwest of England, led by Cardiff University alongside Swansea, Exeter, Bath, and Bristol Universities, the charities Adferiad and Bipolar UK.

Psychotic and bipolar disorders are associated with high disease burden, severely reduced quality of life and premature mortality of between 10-20 years compared to the general population.(48–50) Nevertheless, understanding of these conditions remains relatively limited. Unlike other areas of medicine, psychiatric diagnosis is based exclusively on symptom groups, not yet informed by aetiology or supported by biological markers. Often, symptoms overlap across diagnostic categories and diagnoses change, while individuals with the same diagnosis may have markedly different clinical profiles, treatment responses, and outcomes. This diagnostic heterogeneity, instability, and overlap continue to hinder mental health research and delivery of effective, personalized care.(51–54) To improve outcomes, we must deepen transdiagnostic understanding of aetiology, identify early risk markers, and develop better predictive tools.

The BG Hub has two major research programmes:

1. The Bipolar, Schizophrenia, Psychosis Research Initiative (B-SPRINT) — a recruited cohort study.
2. The Brain and Genomics Hub e-Cohort — a population-level data study using anonymised linked records.

B-SPRINT will recruit a cohort of individuals with schizophrenia, bipolar disorder, and schizoaffective disorder (target N = 600). Data collection will include self-report questionnaires, detailed lifetime symptom interviews, cognitive tasks, biological samples for genomics and biomarkers, MEG and MRI brain imaging, and consent for linkage to routine health and social records for retrospective and longitudinal analyses. Once collected, secure and anonymised data will be made available to other researchers. The BG Hub e-cohort study aims to identify early signs and risk factors and will use de-identified data from the SAIL Databank via the DATAMIND Hub, which securely links health, education, and social care records for people living in Wales.

##### 2.1.3 Study design: the BG Hub *Impact Log*

###### 2.1.3.1 BG Hub LEI

The *Impact Log* has been co-produced within a broader framework of LEI aims and activities.

**Figure 1:**
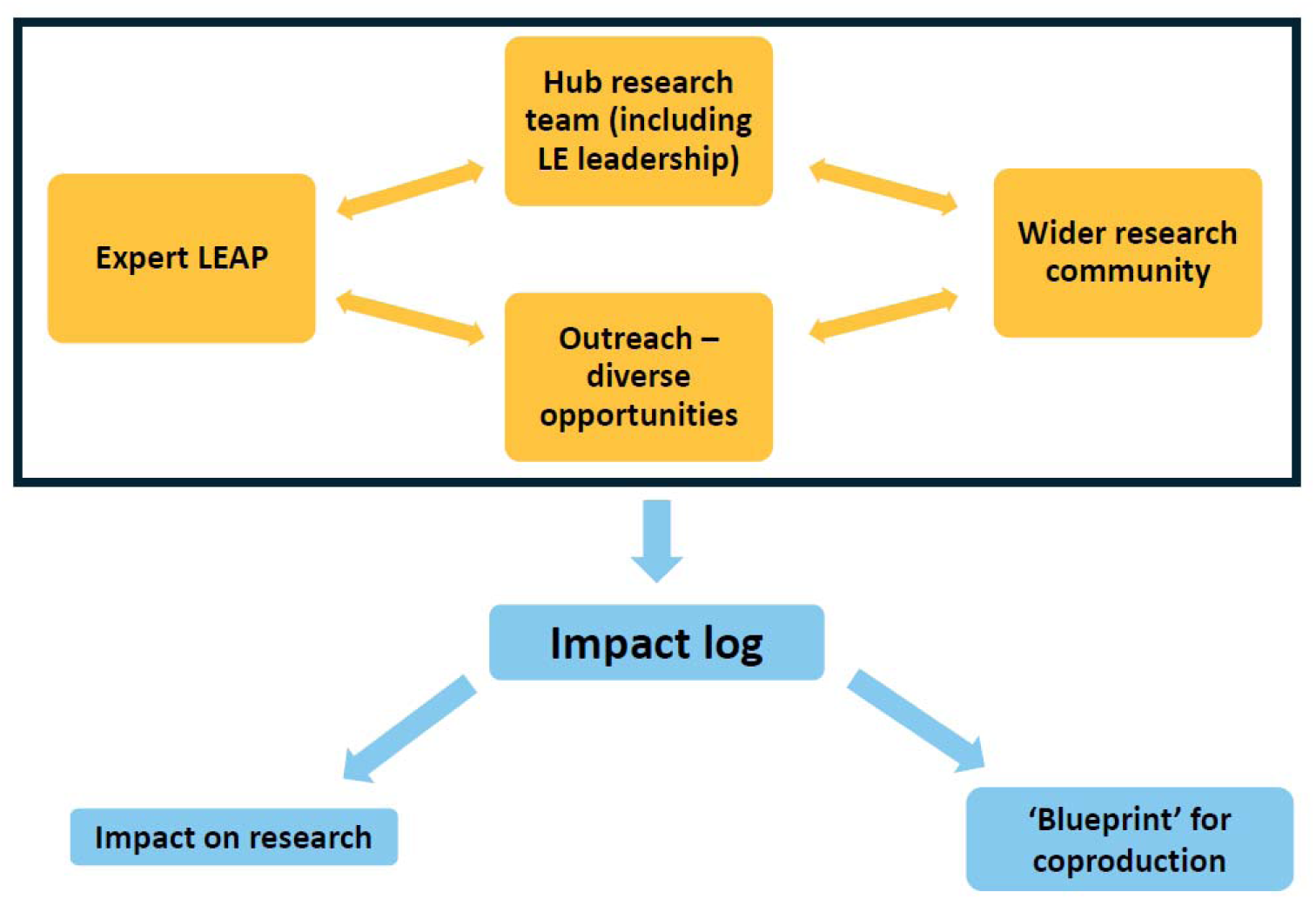
BG Hub LEI and *Impact Log* Structure.

The BG Hub is implementing a complex LEI model to overcome certain common barriers to LEI impact. LEI research opportunities are often restricted to a few individuals, with input limited by time, budgets, and assumptions about where LEI may not be feasible or valuable. There is restricted demographic diversity across age, sex, and ethnicity. LE is often defined broadly, and studies often involve people without direct LE of the relevant condition, particularly with severe mental illness.

The BG Hub works with institutional, LE, and charity partners to recruit diverse participants broadly representative of regional demographics, and build a participant “community”, offering LEI opportunities. B-SPRINT consent includes options for consent to being contacted about LEI opportunities: “I agree to let the study team contact me about opportunities to take part in both paid and unpaid lived experience advisory work. This includes opportunities where my lived experience of bipolar, psychosis, schizophrenia or schizoaffective disorder may be helpful in terms of giving advice to the research team in relation to the design, analysis and dissemination of this and future studies”.

We will develop varied and accessible LEI opportunities, aiming to involve a minimum of 120 (20%) diverse study participants in remunerated work. We included “unpaid” opportunities to allow, for example, for future surveys where budgetary and logistical constraints may make payment impractical. However, all substantive LEI will be remunerated in accordance with NIHR guidelines.(55) For LE core research team members, remuneration follows current institutional salaries.

Given the BG Hub complexity, we also include what we have termed “dual expertise”, which refers to people combining LE/“expertise by experience” with research and/or clinical expertise.(56, 57) Our dual expertise researchers bridge the gap between the research and wider community, enhancing inclusive, accessible LEI for people without prior research experience and supporting non LE researchers. There are six to nine dual expertise researchers with LE of bipolar and psychosis within the BG Hub leadership team and its Specialist LEAP (Lived Experience Advisory Panel - for details of LEAP demographics see Authors’ Contributions). There may be occasional membership fluctuations due to changing commitments amongst LEAP members. Career development opportunities and training will be available, with the Specialist LEAP offered opportunities to broaden research experience and other LE advisors offered an introduction to research.

###### 2.1.3.2 Charity partners

Two BG Hub charity partners, Adferiad and Bipolar UK, are represented within Hub leadership, including co-investigators. Bipolar UK provides bipolar-related peer support, education, advocacy and research, with many staff having LE. Adferiad is a member-led charity providing support for people with mental ill health, addiction, and co-occurring complex needs across Wales, with established LE networks. Adferiad supports LEI in research to enhance inclusivity, representativeness, and impact.

#### 2.1.4 *Impact Log* development

##### 2.1.4.1 Initial adaptation of the Impact Log for the BG Hub

The BG Hub *Impact Log* is based on versions designed and used by TG and TK in the multidisciplinary five-year Wellcome Trust-funded Mental Health and Justice project [For more details see Extended Materials 6]. Based on the initial iteration’s success and feasibility, the BG Hub team planned to use an adapted version of the pilot *Impact Log* to gather data on LEI impact. The Impact Log idea and original form was shared with the LEAP and wider BG Hub team. [See Table 3 for a list of initial adaptations made to the Mental Health and Justice Impact Log].

**Table 3:** Adaptations to the Mental Health and Justice Impact Log.

|  |  |
| --- | --- |
| 1 | To ensure that a broad understanding of impact is reached, the impact LEI is understood as impact on study design and delivery, on all people involved in co-producing research, and on the research environment. |
| 2 | To ensure that a full range of perspectives on LE impact are reflected, *Impact Log* data will be collected from everyone involved in any form of LEI activity, including non-LE research team members, the BG Hub LEAP, and LE advisors taking part in individual research activities. |
| 3 | To ensure that people feel able to respond openly and honestly about their views, forms will be anonymous. All identifying information will be removed before analysis so that data is anonymised and non-identifiable. |
| 4 | To ensure data security, *Impact Log* data will be collected and stored in accordance with the BG Hub wider data protection protocols. |
| 5 | To maximise ease and accessibility, data will be collected via a simple electronic form, with paper versions made available if requested, and any data collected on paper transferred into secure data storage by a study team member. |
| 6 | To maximise rigour in governance, the usability of data, and its potential impact, the BG *Impact Log* forms and process will be included in the BG Hub study protocol and Ethics Committee assessment. |
| 7 | To ensure that a mixture of reflective and standardised data is collected, *Impact Log* forms will include a mixture of quantitative (closed) and qualitative (open) questions, amenable to mixed-methods analysis. |

##### 2.1.4.2 Form development

Following initial consultation and piloting, initial versions of the co-designed BG Hub *Impact Log* forms were approved and refined (See Extended Materials 1-3 for the *Impact Log* forms). For the BG Hub there are three forms:

1. The “Specialist LEAP” Form, for LEAP members with combined LE/research expertise, who will regularly contribute, as BG Hub research team members.
2. The “Lived Experience Involvement” Form, for people with LE participating in individual BG Hub involvement activities, with no expectation of specialist research experience.
3. The “Researcher” Form, for BG Hub research team members not working as LE researchers and (unless undisclosed) lacking direct LE.

Initial piloting included the BG Hub Participant Journey Day, a day-long remunerated visit to the B-SPRINT study site during the B-SPRINT project design phase, for eight people with LE, who would fit BG Hub inclusion criteria, but had no prior BG Hub knowledge. They were led through the experience of the planned study process and provided feedback, including a paper version of the pilot LEI *Impact Log* form, with requests for feedback on layout, content, and usability. Answers and feedback showed positive engagement with the *Impact Log* process, which could be comfortably completed within five minutes. Participants also highlighted the importance of providing digital versions.

Following these stages, the current Forms begin with a section requesting consent and a section requesting details of date, topic, and people involved in LEI activity. People are then asked three pairs of questions about opportunities to contribute, adequacy of explanations given, and the extent to which they felt their opinion was listened to. Each pair includes a quantitative question using a Likert scale to gauge extent and a free text question asking for an explanation of this rating. Finally, for people with LE, there are four open questions including reflections on anticipated impact, any impact on well-being, and more general reflections. Researchers are given questions asking about how LE involvement has impacted on the specific aspect of the research process which they brought to LE advisors and the impact of participating in LEI activities on them as researchers. LEAP and research team members engaged with ongoing BG Hub LEI activities on an ongoing basis are assigned an optional unique identifier number to include on their *Impact Log* forms, to allow longitudinal analysis of *Impact Log* data.

#### 2.1.5 Data management and analysis

##### 2.1.5.1 Data collection and storage

Digital versions of the *Impact Log* Forms were created using REDCap software, with all *Impact Log* data stored securely on Cardiff University servers, following data security protocols and B-SPRINT ethics permissions. Data is only accessible to members of the BG Hub research team with permissions and an appropriate institutional login. Any Specialist LEAP members or Charity partner representatives choosing to be involved in *Impact Log* data analysis will be trained in data governance and given data access via secure Cardiff University servers.

*Impact Log* forms will be distributed to anyone involved in BG Hub LEI activities after completing the activity, usually via a secure link or QR code. Initial consultation has shown that a digital form is generally preferred. However, it will be made clear that written forms are available if preferred. For researchers and LEAP members, forms will be circulated after initial actions have been implemented, to facilitate reflection on any implemented or planned changes resulting from this LEI activity. *Impact Log*s will be distributed and completion tracked by TG, the BG Hub LE lead. TG will not complete *Impact Log* forms, to mitigate risk of bias, given that TG will be viewing and tracking all forms and leading on *Impact Log* development and analysis.

##### 2.1.5.2 Data analysis

Two types of data analysis are planned. All *Impact Log* data collected through the BG Hub will be analysed in the final year, including longitudinal analysis of “Specialist LEAP” and “Researcher” forms, if digital identifiers have been included. There will also be iterative informal and formal analysis of *Impact Log* data at various stages. Results of early analysis will be used to facilitate research team reflections on how the BG Hub LEI process is working and evolving, and whether any adaptations to LEI or the *Impact Log* should be considered. A key part of analysis will be evaluation of the *Impact Log* tool itself, examining how well it is working, recording dates and contexts for circulating *Impact Log* forms, monitoring response rates and ease of use, recording any modifications made during the course of the project, and surveying users of the *Impact Log* at various stages.

Given that the *Impact Log* tool is a novel tool which does not incorporate established outcome measures, analysis of *Impact Log* data will be an exploratory process. Analysis will focus on assessing impact across the three domains identified in our aims: impact on research design, delivery, and outcomes; systemic impact on research processes and environment; interpersonal and cultural impact within the research community. To achieve this, *Impact Log* data will be considered in relation to the wider research context of LEI activities being evaluated. Analysis will be led by the BG Hub LE lead, with assistance from specialist LEAP and other BG Hub research team members. To ensure greater inclusivity and representativeness, we also aim to involve LE advisors from the wider study community in analysis and dissemination, following a framework to be developed by the specialist LEAP.

Data will be extracted into Excel for cleaning and analysis, given Excel’s suitability for mixed-methods analysis of non-complex data. Using Excel also allows for maximal accessibility for LEI in data analysis. Finally, a qualitative analysis tool, such as NVivo, may be used, and access to NVivo software will be provided for all researchers involved.

As the forms combine quantitative and qualitative questions, it is anticipated that we will analyse quantitative data descriptively using standard statistical methods, and qualitative data using thematic analysis. Quantitative and qualitative data will be analysed separately and then brought together to contextualise and explore patterns observed in the numerical responses. Where appropriate we will use regression models to analyse longitudinal changes in ratings, using statistical software decided in collaboration with the LEAP, and narrative synthesis to describe how themes or experiences evolve.

#### 2.1.6 Dissemination

The BG Hub *Impact Log* has two key dissemination objectives: to contribute to ongoing development and dissemination of LEI activities and their impact within the BG Hub; and, to inform design and delivery of other mental health research, and the training of mental health researchers and clinicians. Our hope is that the *Impact Log* and surrounding LEI framework can work as an adaptable blueprint for complex and impactful LE involvement in research.

*Impact Log* materials and evolving analysis will be shared internally within the BG Hub to support ongoing use and development of consistent LEI across workstreams. *Impact Log* materials, forms and guidance will also be made publicly available, enabling replication and adaptation elsewhere. Along with *Impact Log* data, study documentation (e.g., protocols, consent templates, and other participant-facing materials) will be hosted on DATAMIND, the designated data resource sharing platform of the UKRI Mental Health Platform. This data will be accessible via a data access committee to bona fide researchers. The availability of the data on DATAMIND promotes transparency and reproducibility and may be particularly useful for early career researchers seeking to conduct similar research.

*Impact Log* findings will be communicated in multiple formats and tailored to different audiences. Formal academic dissemination will include peer-reviewed publications and conference presentations. For applied settings, findings will be integrated into CPD activities such as internal training sessions or knowledge-exchange events for clinicians, researchers and LE experts.

Informal and public-facing dissemination will focus on accessibility and engagement. Key findings will be summarised through infographics, plain-language summaries, social media posts, and short animated or recorded videos designed for lay audiences. These formats will be developed collaboratively with LEAP members to ensure alignment with LE communication preferences and priorities.

Overall, the methods and data generated by the *Impact Log* initiative have broad application scope and significant potential to be useful for clinicians, researchers, experts by experience, lay audiences, and ethics committees. Our dissemination strategy maximises reach and utility of the *Impact Log* methodology and findings. By sharing materials openly and in multiple formats, we aim to support public understanding, enhance research and clinical capacity, and contribute to the evolving definition of meaningful LEI impact in research.

#### 2.1.7 Lived experience perspectives on the *Impact Log* design and dissemination process

To understand more about LE experiences of participating in this project, a Specialist LEAP member, JK, created a short anonymous survey and conducted analysis, asking Specialist LEAP members about experiences of contributing to the *Impact Log* design and dissemination process, and reflections on its potential significance. Four main themes emerged: the importance of collaboration; the value of integration within the research team; the *Impact Log*’s potential as a methodological resource; the value of the *Impact Log* for increasing transparency in research and demonstrating the value of LEI (For Themes and selected illustrative quotes, see Table 4. For full results see Extended Materials 7).

**Table 4:** Themes and selected illustrative quotes for each theme Abbreviations: LEAP = Lived Experience Advisory Panel

| Theme: | Selected illustrative quotes: |
| --- | --- |
| The importance of collaboration | Bringing in lived experience perspectives adds dimensions and ideas to the research that might otherwise be overlooked if researchers worked in isolation. That diversity of perspective not only enriches the process but also makes any tool, or team, or project, stronger and more impactful. |
|  | The LEAP meetings have been a great opportunity to hear the views and experiences of other researchers with lived experience and I think our voices together are really powerful and add unique insights into the project. |
| The value of integration with the research team | Working alongside the academic team has really helped equalise that power imbalance that is inherent in researcher-LEAP relationships. |
|  | But the LEAP and *Impact Log* publication team are well integrated into the wider study team and making what is recognised to be an immensely important contribution to the work within the project. |
|  | Co-production initiatives are so often hampered by tokenism, power imbalances, and failures to be representative. But the LEAP and *Impact Log* publication team are well integrated into the wider study team and making what is recognised to be an immensely important contribution to the work within the project. |
| The *Impact Log* as a methodological resource | I am hopeful that the IL [*Impact Log*] tool can provide a tool-kit and framework which others will be able to adapt and use within their research, so that we move forward in making co-production a more methodologically developed component of research, as well as being a tool which recognises the interpersonal components of co-production work and the difference this can make to everyone involved. |
|  | I think the *Impact Log* evaluation tool is going to be beneficial in helping us operationalise something that is inherently subjective and often difficult to convey. |
|  | This is relevant for all stakeholders of brain & genomics hub project - including funding bodies. In a larger context, the *Impact Log* evaluation tool can be utilized across the mental health research field. |
| Increasing transparency/demonstrating the value of co-production with the *Impact Log* | This kind of evaluation is really needed in LEAP advisory work, because it allows us to move beyond tokenistic involvement of those with lived experience and identify where lived experience input has the biggest impact, and where it could be strengthened. In turn, that makes the process more transparent, accountable, and ultimately more meaningful—both for researchers and for the communities the research is meant to serve. |
|  | Being able to measure and demonstrate this impact is a really important part of co-designing research and is an important part of the process of participating in research as someone with lived experience. |

LEAP members emphasised the importance of collaboration within the LEAP and wider research team. Key themes were the potential of LE voices to enrich the research process; the power of including diverse voices; and the learning that takes place during co-production. Another key theme was the value of LE integration into the wider study team, and how integration facilitates recognition and incorporation of LE contributions with potential to mitigate tokenism and power imbalances in LEI.

Another theme was the *Impact Log*’s potential as a methodological resource. LEAP members emphasised its potential utility across different stages of research and its potential to capture and operationalise the inherently subjective and nuanced process of LEI. They also highlighted its potential utility beyond the BG Hub and how formal recording and evaluation of LE involvement in mental health and wider medical research might help embed LEI as a core methodological component. This might extend to other stakeholders including, for example, funding bodies.

Finally, LEAP members emphasised the *Impact Log*’s potential to increase transparency and accountability within research and demonstrate LEI’s value. They suggested that the *Impact Log* tool might help researchers reduce tokenism and identify areas where LEI has greatest impact and areas where LEI could be improved and strengthened. The *Impact Log* was viewed as offering mutual benefits to the research project and, also importantly, to LE researchers by enabling them to see the impact of their involvement, making the research process more beneficial and meaningful and supporting their own development.

The implications of the LE perspective survey responses can be summarised as follows. First, the Impact Log tool has potential value for all research stakeholders and could help to establish LEI as a key research component, presenting a framework also usable, for example, by funding and policy institutions. Second, working explicitly to increase integration and significance of LE roles within research projects, rather than framing LEI as supplementary, will increase LEI’s meaningfulness and impact for everyone involved. Third, a tool to evaluate impact and identify any problems in the LEI process will promote transparency and accountability and help identify and address areas where LEI is not working well, potentially mitigating tokenism and sidelining of LE contributions.

## 3 Discussion

This protocol outlines the background, development and implementation plans for the BG Hub *Impact Log* tool, together with its surrounding LEI framework and research context. This tool will allow us to record and evaluate LEI impact across three research domains: design and delivery; systems and process; interpersonal and environmental. The *Impact Log* and LEI framework also provides a replicable tool, easily adaptable for use elsewhere in medical research. We must emphasise that the *Impact Log* remains a novel tool. Both design and analysis will involve iterative development, as we observe the tool and framework performing within the BG Hub research context.

### 3.1 Strengths and limitations

A key strength of the *Impact Log* and LEI framework is explicit design to address widely recognised challenges in LEI, particularly tokenism and gaps in structural integration, diversity, and acknowledgement of LEI’s complex and multifaceted potential. LEI is fully integrated, with robust plans to ensure LE perspectives inform all research stages. The core study team, including senior leadership, includes people with LE of the conditions being researched. The strength and value of this collaborative integration is already evidenced in our survey of LE perspectives on *Impact Log* design and dissemination (2.1.7).

LE leads, specialist LEAP members and other LE advisors have already had significant impact on major study design decisions and involvement in early dissemination including the study launch, with structures in place to ensure these contributions continue. Systemic adaptation and structural integration are further achieved by ensuring LE contributions are swiftly and adequately remunerated, and training, support, and resources provided when needed. The BG hub is underpinned by a dedication to demographic and health inclusivity and diversity. Our specialist LEAP exemplifies this approach, with members from minoritised ethnicities, varied ages, gender and sexual minorities, and disabilities (see Authors’ Contributions below for further details on LEAP demographics). This produces nuances within discussion which would otherwise be neglected. Moreover, the broader LEI framework is designed to dedicate resources to ensuring diversity and inclusivity are also embedded in wider LEI.

To our knowledge, the *Impact Log* and its LEI framework are highly innovative in many regards. A major strength is high potential for adaptation and implementation elsewhere, thereby contributing towards methodological development in LEI.

There are some potential limitations, however, relating to both the BG Hub context and wider implementation. Our *Impact Log* framework depends on resource availability, including funding, people, and time, which may be challenging elsewhere, particularly if LEI resources are limited during individual stages or throughout a project. Successful implementation of the *Impact Log* will require sustained commitment to LEI and evaluation throughout research delivery. This may be difficult, even within the BG Hub team’s deep commitment to LEI, particularly when time is limited. Such challenges would be exacerbated by any team power imbalances leaving LE team members with less power to press for prioritisation of LEI.

Ideally, implementation also requires people with LE and research experience embedded in the leadership team. This might not always be possible. TG and TW had salaried positions supporting LEI in developing the BG Hub application, enabling incorporation of the *Impact Log* and LEI framework into study design from the outset. Generally, however, a well-known major barrier to successful LEI is lack of funding supporting LEI in initial project development and funding applications, which could affect *Impact Log* integration.

The *Impact Log* framework involves continuing input of LE lead/s and other LE research team members. Ongoing health challenges could therefore present difficulties, particularly in research on severe and chronic illness. Ideally, contingency plans should ensure that LEI and impact evaluation can continue during any research team absences or reduced work capacity. Nevertheless, resourcing this may be difficult and LE researchers may well feel uncomfortable disclosing impact of health on work, if concerned about negative judgements about their research capacity.

Many factors might contribute to risk of bias in *Impact Log* responses submitted by LE and non-LE researchers, especially if people have concerns about anonymity. Although forms will be analysed anonymously, respondents are likely to be identifiable to the study team member/s responsible for collation and analysis. The potential for identification may be particularly high in smaller studies, with smaller LEAPs or fewer LE advisors, and in longer studies where LE and non-LE team members submit multiple *Impact Log* forms, especially if linked by an *Impact Log* ID number. Anonymity-based concerns may constrain respondents’ honesty, particularly for negative opinions, if people have concerns about causing offence or impacting negatively on the project. Conversely, negative views, for example, about the research model of a particular project, could potentially lead to negative bias in *Impact Log* responses. These risks may be slightly mitigated by emphasising the importance of honest disclosure, balance, and the fact that analysis of forms is collective with identifying details removed. Moreover, if people feel confident in their integration and status within the study team, this may increase their confidence in providing honest answers, even if critical. Nevertheless, the potential for bias must be recognised and considered throughout.

### 3.2 Future directions

The *Impact Log* will be used throughout BG Hub research to collect data informing ongoing tool development. We also hope our protocol will provide sufficient information and materials for wide dissemination, adaptation, and timely use of the *Impact Log*, and BG Hub team members already have plans to begin its implementation elsewhere.

The *Impact Log* tool and LEI framework involve many factors relevant for ongoing LEI discussions and methodological development. Alongside its development and dissemination within the BG Hub, much work remains for BG Hub researchers and others, exploring wider methodological developments and creating future research projects enabling their implementation and evaluation. Finally, we hope the *Impact Log* project might support advocacy for promotion and integration of impactful, representative, and equitable LEI in medical research and practice.

#### 3.2.1 Creating an Impact Log and LEI community – please keep in touch

To develop the *Impact Log* further, we would like to know about people’s views and possible adoption of this tool, building more collective understanding of its adaptability for different contexts. If you are considering using the *Impact Log*, we would be very grateful if you might consider informing the corresponding author, TG, by email. Currently, there is no formal feedback process, but we would like to start creating a network allowing for future possibilities.

## Supporting information

Extended materials Gergel et al. Impact Log Protocol

## Data Availability

Datasets generated and analysed in preparing this article are available in Supplementary Materials. These comprise the full set of responses for the survey Lived experience perspectives on the Impact Log design and dissemination process (Supplementary Materials 6) and a table of themes and reference list for the narrative review (Supplementary Materials 5). Given its length, the data extraction table for the narrative review has not been included in the Supplementary Materials. However, it will be made available to researchers via the corresponding author if contacted.
The Impact Log data collected during the BG Hub will be disseminated in accordance with the plans described above under Dissemination. Given that the Impact Log forms will gather potentially detailed qualitative data, full publication of the Impact Log forms dataset will not be possible. However, within the dissemination process, there will be consultation within the BG Hub research team and LE advisors from the wider Hub community about the publication of sets of collated responses to particular questions, with identifying details removed.

## Abbreviations

LE: Lived Experience.
LEI: Lived Experience Involvement
BG Hub: Brain and Genomics Hub.
LEAP: Lived Experience Advisory Panel.

## Data availability

The *Impact Log* data collected during the BG Hub will be disseminated in accordance with the plans described above under “Dissemination”. Given that the *Impact Log* forms will gather potentially detailed qualitative data, full publication of the *Impact Log* forms dataset will not be possible. However, within the dissemination process, there will be consultation within the BG Hub research team and LE advisors from the wider Hub community about the publication of sets of collated responses to particular questions, with identifying details removed.

## Extended Data

Datasets generated and analysed in preparing this article are available in Extended Materials (https://doi.org/10.17605/OSF.IO/NRCFY). These comprise the full set of responses for the survey “Lived experience perspectives on the *Impact Log* design and dissemination process” (Extended Materials 7) and a table of themes and reference list for the narrative review (Extended Materials 5.3/4). Given its length, the data extraction table for the narrative review has not been included in the Supplementary Materials. However, it will be made available to researchers via the corresponding author if contacted.

## Ethics approval and consent to participate

B-SPRINT, including the *Impact Log* has received ethical approval from Wales REC 7 (reference: 25/WA/0027).

## Authors’ contributions

All authors have contributed to the development of the Impact Log and the preparation of this manuscript. In addition: TG, TW, CE, EV, LG, HG, JK, OD are involved in the specialist dual expertise LEAP (lived experience advisory panel). TG and TW led on development and implementation of the LEI strategy and impact log for the Brain and Genomics Hub, and on the Bipolar UK partnership. CH and JHe led on the Adferiad partnership. TG and TK developed the initial version of the impact log. EV, LG, CE, HG, TG, and JHe worked on the narrative review. JK designed and analysed the survey within the protocol “Lived experience perspectives on IL design and design dissemination process”. OD and CH led on developing the dissemination plans for publication. TW and CM led on the form creation and data management plans. TG, CB, GD, SL, JW, NH, AJ, IJ, JHa, CH, Jhe, TW have all contributed to the development and implementation of the Brain and Genomics Hub LEI strategy and impact log, in their capacity as members of the central research team for the Brain and Genomics Hub. [See Table 1 for author disclosed information on mental health LE and professional experience.]

### Hub LE researcher / specialist LEAP demographic details

All eight BG Hub LE researchers provided some basic demographic information, with the agreement that this would be collated and described to ensure anonymity and options to choose not to answer particular questions. Four LE researchers (50%) are aged between 25- 34; with three between 35-44 and one researcher aged 45-54. Gender of LE researchers included male, female, trans-female, and non-binary. Disclosed sexuality was almost evenly split between researchers identifying as heterosexual/straight and bisexual. Half the team identified as white, with three other ethnic groups represented. Three researchers identified as having no religion, while five distinct religions/beliefs were named by the remaining researchers. Finally, under ‘additional information’ one researcher identified LE of mood disorder not captured in the Authors’ Contributions table.

## Competing interests

TG’s work is supported by the following awards: NIHR167361 (Home-based transcranial direct current stimulation in bipolar depression: a randomised, double-blind, placebo-controlled trial); NIHR132773 (Aripiprazole/ sertraline combination: a non-inferiority design in comparison with quetiapine in bipolar depression. An open label randomised controlled trial); NIHR Mental Health Translational Research Collaboration. OD receives consulting fees from McGill University (DIALOG) and the Feinstein Institutes (VITAL); TK has received consultancy fees from Bristol Myers Squibb and Boehringer Ingelheim; has received research grants from Takeda Pharmaceuticals and has received research grants and acted as a consultant for Akrivia Health for work unrelated to the current manuscript.

## Grant Information

The work of all authors on this manuscript is supported by funding from the Medical Research Council (MRC) to the Brain and Genomics Hub of the Mental Health Platform (Grant No. MR/Z503745/1). TG and TK also received funding for the initial development of the Impact Log from the Wellcome Trust, UK (grant number 203376/Z/16/Z), as part of the Mental Health and Justice project. The Brain and Genomics Hub is also supported by The National Centre for Mental Health, a collaboration between Cardiff, Swansea, and Bangor universities funded by the Welsh Government through Health and Care Research Wales.

AJ receives funding from the MRC for DATAMIND (MR/Z504816/1) and the National Centre for Suicide Prevention and Self-Harm Research (Health and Care Research Wales); JHa is supported by the Hodge Foundation and The Waterloo Foundation; TG receives and TW received salary from Bipolar UK (until 30^th^ August 2025). CH and JHe receives salary from Adferiad.

## Notes

### Competing Interest Statement

TGs work is supported by the following awards: NIHR167361 (Home-based transcranial direct current stimulation in bipolar depression: a randomised, double-blind, placebo-controlled trial); NIHR132773 (Aripiprazole/ sertraline combination: a non-inferiority design in comparison with quetiapine in bipolar depression. An open label randomised controlled trial). OD receives consulting fees from McGill University (DIALOG) and the Feinstein Institutes (VITAL); TK has received consultancy fees from Bristol Myers Squibb and Boehringer Ingelheim; has received research grants from Takeda Pharmaceuticals and has received research grants and acted as a consultant for Akrivia Health for work unrelated to the current manuscript.

### Author Declarations

B-SPRINT, including the Impact Log has received ethical approval from Wales REC 7 (reference: 25/WA/0027).

### Summary of Updates

The text and materials have been substantively revised to help with readability and condense, and also to include the latest updates of the impact log forms, including: 1. Revisions of the impact log forms to increase clarity and request more information. 2. Some changes to terminology, including more general use of term lived experience involvement rather than co-production as a general term. 3. Condensing of the initial brief narrative literature review, to help with brevity and clarity. This is now included as part of the introductory section, with methodological details moved to the Extended Materials section. 4. Condensing / moving of details relating to the initial Mental Health and Justice Impact Log Form to Extended Materials.

