## Extended materials Gergel et al. Impact Log Protocol for "A tool to evaluate the impact of lived experience involvement in research: the Brain and Genomics Hub *Impact Log* protocol"

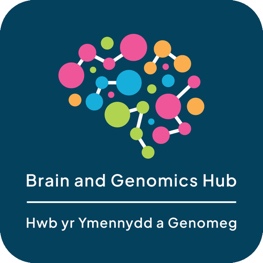

### 1. Lived Experience Involvement IMPACT Log Form

**Introduction**

We would like to ask you some questions to help us understand more about the impact that lived experience involvement can have on research. You have recently taken part in a lived experience involvement activity for the Brain and Genomics Hub and helped with research as someone with lived experience. We will ask you a few questions about your experience. This will include questions about what was discussed, how the group worked together, and your experience of taking part.

This form should take no more than 5 minutes to complete. The answers you give will be stored securely and kept anonymous. The Brain and Genomics Hub research team will be analysing the information provided by you and others in order to:

1. Help us understand more about how lived experience involvement in the Hub is working, and whether there are things we need to improve.
2. Help us provide information for other researchers and the wider community about the difference that lived experience involvement can make in research, and what areas need to be worked on more.

**Data access and privacy**

The information you supply will be anonymously stored on Cardiff University secure servers in line with Brain and Genomics Hub ethical approvals (REC reference: 25/WA/0027). Your data will only be accessed by approved members of the Brain and Genomics Hub research team and approved collaborators, in accordance with the Hub's data access governance processes.

**Please read each statement below and indicate whether you agree.**

**You are free to withdraw from this study at any time without giving a reason. If you no longer wish to have your data stored, please contact the study team at****.**

| 1) Have you read and understood the information presented above, read the privacy notice, and consent to Cardiff University collecting your data? | - Yes - No |
| --- | --- |
| 2) Do you consent to your anonymised data being shared with researchers within the wider Brain and Genomics Hub for purposes of analysis and dissemination? | - Yes - No |
| 3) Do you consent to your anonymised data being used in reports and publications that may arise from this work? | - Yes - No |

The questions we will now ask are being used to understand more about how your involvement as someone with lived experience has impacted on the lived experience involvement activity you took part in. We are doing this to record and evaluate how lived experience involvement is impacting on research within the Brain and Genomics Hub and to understand more about whether any changes and improvements could be made**.**

**All questions are optional – the text questions are there for you to give more detail if you wish.**

**If there are any questions you do not want to answer or cannot answer at all or fully, don’t worry and just move onto the next question.**

4) Date of the lived experience involvement activity you took part in?
5) Who did you meet with when you took part in this lived experience involvement activity?
6) What questions or topics were the main focus of this lived experience involvement activity?

**Did you feel you were given the chance to contribute to this lived experience involvement activity?**

- If you agree that you did have the chance to contribute, please tick either 'strongly agree' or 'agree'.
- If you feel did not have the chance to contribute, please tick either 'disagree' or 'strongly disagree'.
- If you do not feel strongly that you agree or disagree, please tick 'neutral'.

7) I felt I was given the chance to contribute to this lived experience involvement activity.

| *Strongly disagree* | *Disagree* | *Neutral* | *Agree* | *Strongly agree* |
| --- | --- | --- | --- | --- |

8) Can you explain why you gave this answer about whether you felt you had the chance to contribute?

**Do you think the research questions or topics for this lived experience involvement activity were explained well?**

- If you agree that the research questions or topics were explained well, please tick either 'strongly agree' or 'agree'.
- If you feel that the research questions or topics and question(s) were not explained well , you should tick either 'disagree' or 'strongly disagree'.
- If you do not feel strongly that you agree or disagree, please tick 'neutral'.

9) I felt the research questions or topics and question(s) for this lived experience involvement activity were explained well

| *Strongly disagree* | *Disagree* | *Neutral* | *Agree* | *Strongly agree* |
| --- | --- | --- | --- | --- |

10) Can you explain why you gave this answer about whether you felt research questions or topics were explained well?

**Did you feel that your opinion was listened to during this lived experience involvement activity?**

- If you agree that your opinion was listened to, please tick either 'strongly agree' or 'agree'.
- If you feel that your opinion was not listened to, please tick either 'disagree' or 'strongly disagree'.
- If you do not feel strongly that you agree or disagree, please tick 'neutral'.

11) I felt that my opinion was listened to

| *Strongly disagree* | *Disagree* | *Neutral* | *Agree* | *Strongly agree* |
| --- | --- | --- | --- | --- |

12) Can you explain why you gave this answer about whether you felt that your opinion was listened to?
13) What do you hope will be acted on or changed because of your contributions to this lived experience involvement activity?
14) Has taking part in this lived experience involvement activity had any impact on your well-being?
15) Is there anything else you would like to say, as someone with lived experience, about your experience of taking part in this lived experience involvement activity?

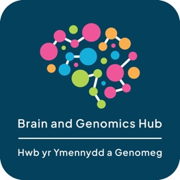

### 2. Brain and Genomics Hub Specialist LEAP IMPACT Log Form

**Introduction**

We would like to ask you some questions to help us understand more about the impact that lived experience involvement can have on research. You have recently taken part in a Specialist Lived Experience Advisory Panel (LEAP) activity for the Brain and Genomics Hub. We will ask you some questions about your experience of contributing to research as someone with both lived experience and research or clinical experience. This will include questions about what was discussed, how the group worked together, and your experience of taking part.

This form should take no more than 5 minutes to complete. The answers you give will be stored securely and kept anonymous. The Brain and Genomics Hub research team will be analysing the information provided by you and others in order to:

3. Help us understand more about how lived experience involvement in the Hub is working, and whether there are things we need to improve.
4. Help us provide information for other researchers and the wider community about the difference that lived experience involvement can make in research, and what areas could be developed or improved.

**Data access and privacy**

The information you supply will be anonymously stored on Cardiff University secure servers in line with Brain and Genomics Hub ethical approvals (REC reference: 25/WA/0027). Your data will only be accessed by approved members of the Brain and Genomics Hub research team and approved collaborators, in accordance with the Hub's data access governance processes.

**Please read each statement below and indicate whether you agree.**

**We are interested in how your responses to the LEAP Impact Log Forms have progressed over the project.** To do this, we have assigned you a **personal identifier number (Impact Log ID)** so we can combine all the information you have provided. If you give us permission to do this, please input your Impact Log ID number below (or leave blank if you do not wish to have your data combined).

**You are free to withdraw from this study at any time without giving a reason. If you no longer wish to have your data stored, please contact the study team at****.**

| 1) Impact Log ID: |  |  |
| --- | --- | --- |
| 2) Have you read and understood the information presented above, read the privacy notice, and consent to Cardiff University collecting your data? | | - Yes - No |
| 3) Do you consent to your anonymized data being shared with researchers within the wider Brain and Genomics Hub for purposes of analysis and dissemination? | | - Yes - No |
| 4) Do you consent to your anonymized data being used in reports and publications that may arise from this work? | | - Yes - No |

The questions we will now ask are being used to understand more about how your involvement as someone with lived experience has impacted on the specialist LEAP activity you took part in. We are doing this to record and evaluate how lived experience involvement is impacting on research within the Brain and Genomics Hub and understand more about whether any changes and improvements could be made.

**All questions are optional – the text questions are there for you to give more detail if you wish.**

**If there are any questions you do not want to answer or cannot answer at all or fully, don’t worry and just move onto the next question.**

5) Date of the lived experience involvement activity you took part in?
6) Who did you meet with when you took part in this lived experience involvement activity?
7) What questions or topics were the main focus of this lived experience involvement activity?

**Did you feel you were given the chance to contribute to this lived experience involvement activity?**

- If you agree that you did have the chance to contribute, please tick either 'strongly agree' or 'agree'.
- If you feel did not have the chance to contribute, please tick either 'disagree' or 'strongly disagree'.
- If you do not feel strongly that you agree or disagree, please tick 'neutral'.

8) I felt I was given the chance to contribute to this lived experience involvement activity.

| *Strongly disagree* | *Disagree* | *Neutral* | *Agree* | *Strongly agree* |
| --- | --- | --- | --- | --- |

9) Can you explain why you gave this answer about whether you felt you had the chance to contribute?

**Do you think the research questions or topics for this lived experience involvement activity were explained well?**

- If you agree that the research questions or topics were explained well, please tick either 'strongly agree' or 'agree'.
- If you feel that the research questions or topics and question(s) were not explained well, you should tick either 'disagree' or 'strongly disagree'.
- If you do not feel strongly that you agree or disagree, please tick 'neutral'.

10) I felt the research questions or topics and question(s) for this lived experience involvement activity were explained well

| *Strongly disagree* | *Disagree* | *Neutral* | *Agree* | *Strongly agree* |
| --- | --- | --- | --- | --- |

11) Can you explain why you gave this answer about whether you felt research questions or topics were explained well?

**Did you feel that your opinion was listened to during this lived experience involvement activity?**

- If you agree that your opinion was listened to, please tick either 'strongly agree' or 'agree'.
- If you feel that your opinion was not listened to, please tick either 'disagree' or 'strongly disagree'.
- If you do not feel strongly that you agree or disagree, please tick 'neutral'.

12) I felt that my opinion was listened to

| *Strongly disagree* | *Disagree* | *Neutral* | *Agree* | *Strongly agree* |
| --- | --- | --- | --- | --- |

13) Can you explain why you gave this answer about whether you felt that your opinion was listened to?
14) What do you hope will be acted on or changed because of your contributions to this lived experience involvement activity?
15) Has taking part in this lived experience involvement activity had any impact on your well-being?
16) Is there anything that you would like to say about the experience of taking part in lived experience involvement work as someone who has both lived experience and experience of mental health research or clinical work?
17) Is there anything else you would like to say, as someone with lived experience, about your experience of taking part in this lived experience involvement activity?

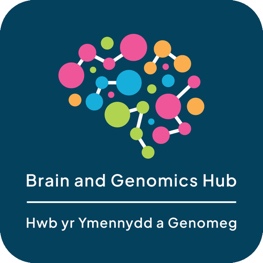

### 3. Researcher IMPACT Log Form

**Introduction**

We would like to ask you some questions to help us understand more about the impact that lived experience involvement can have on research. You have recently taken part, as a member of the research team, in a lived experience involvement activity for the Brain and Genomics Hub. We will ask you a few questions about your experience of working with people with lived experience on research activities within the Brain and Genomics Hub. This will include questions about what was discussed, how the group worked together, and your experience of taking part.

This form should take no more than 5 minutes to complete. The Brain and Genomics Hub research team will be analysing the information provided by you and others in order to:

5. Help understand more about how lived experience involvement in the Hub is working, and whether there are things we need to develop or improve.
6. Help us provide information for other researchers and the wider community about the difference that lived experience involvement can make in research, and what areas require additional work and focus.

**Data access and privacy**

The information you supply will be anonymously stored on Cardiff University secure servers in line with Brain and Genomics Hub ethical approvals (REC reference: 25/WA/0027). Your data will only be accessed by approved members of the Brain and Genomics Hub research team and approved collaborators, in accordance with the Hub's data access governance processes.

**Please read each statement below and indicate whether you agree.**

**We are interested in how your responses to the Impact Log Forms have progressed over the project.** To do this, we have assigned you a **personal identifier (Impact Log ID)** in order for us to collate all the information you have provided. If you give us permission to do this, please input your Impact Log ID below (or leave blank if you do not wish to have your data collated).

**You are free to withdraw from this study at any time without giving a reason. If you no longer wish to have your data stored, please contact the study team at****.**

| 1) Impact Log ID: |  |  |
| --- | --- | --- |
| 2) Have you read and understood the information presented above, read the privacy notice, and do you consent to Cardiff University collecting your data? | | - Yes - No |
| 3) Do you consent to your anonymized data being shared with researchers within the wider Brain and Genomics Hub for purposes of analysis and dissemination? | | - Yes - No |
| 4) Do you consent to your anonymized data being used in reports and publications that may arise from this work? | | - Yes - No |

The questions we will now ask are being used to understand more about how lived experience involvement has impacted on this research activity. We are doing this to record and evaluate how lived experience involvement is impacting on research within the Brain and Genomics Hub and to understand more about whether any changes and improvements could be made.

**All questions are optional – the text questions are there for you to give more detail if you wish.**

**If there are any questions you do not want to answer or cannot answer at all or fully, don’t worry and just move onto the next question.**

5) Date of the lived experience involvement activity you took part in?
6) Who did you meet with when you took part in this lived experience involvement activity?
7) What questions or topics were the main focus of this lived experience involvement activity? (if you were the research team member presenting the main question/topic for discussion, please can you state this).
8) How do you think that lived experience involvement has impacted on the specific research issues or topics which were the focus of this lived experience involvement activity
9) Do you anticipate that there will be additional future impact?

**Did you feel you were given the chance to contribute to this lived experience involvement activity?**

- If you agree that you did have the chance to contribute, please tick either 'strongly agree' or 'agree'.
- If you feel did not have the chance to contribute, please tick either 'disagree' or 'strongly disagree'.
- If you do not feel strongly that you agree or disagree, please tick 'neutral'.

10) I felt I was given the chance to contribute to this lived experience involvement activity.

| *Strongly disagree* | *Disagree* | *Neutral* | *Agree* | *Strongly agree* |
| --- | --- | --- | --- | --- |

11) Can you explain why you gave this answer about whether you felt you had the chance to contribute?

**From your perspective as a researcher, do you think the research questions or topics for this lived experience involvement activity were explained well to the people with lived experience who were asked to be involved?**

- If you agree that the research questions or topics were explained well, please tick either 'strongly agree' or 'agree'.
- If you feel that the research questions or topics and question(s) were not explained well enough, you should tick either 'disagree' or 'strongly disagree'.
- If you do not feel strongly that you agree or disagree, please tick 'neutral'.

12) I felt the research questions or topics and question(s) for this lived experience involvement activity were explained well to the lived experience contributors.

| *Strongly disagree* | *Disagree* | *Neutral* | *Agree* | *Strongly agree* |
| --- | --- | --- | --- | --- |

13) Can you explain why you gave this answer about whether you felt research questions or topics were explained well?

**Did you feel that the research questions raised by the research team during this lived experience involvement activity were focused on enough?**

- If you agree that there was sufficient focus on the research questions raised by the research team, please tick either 'strongly agree' or 'agree'.
- If you feel that there was insufficient focus on the research questions raised by the research team, please tick either 'disagree' or 'strongly disagree'.
- If you do not feel strongly that you agree or disagree, please tick 'neutral'.

14) I felt that that there was sufficient focus on the research questions raised by the research team

| *Strongly disagree* | *Disagree* | *Neutral* | *Agree* | *Strongly agree* |
| --- | --- | --- | --- | --- |

15) Can you explain why you gave this answer about whether you felt that that there was sufficient focus on the research questions raised by the research team?
16) What impact (if any) has taking part in this research activity involving people with lived experience had on your understanding, beliefs, and priorities about severe mental health conditions?
17) What general impact (if any) has taking part in this research activity involving people with lived experience had on you as a researcher?
18) Is there anything else you would like to say about your experience, as a research team member, of being part of this lived experience involvement activity, that has not been mentioned above?

### 4. GRIPP2 long form for reporting Lived Experience Involvement (Guidance for Reporting Involvement of Patients and the Public)

| **Section and topic** | **Item** | **Reported in Section** |
| --- | --- | --- |
| Section 1: Abstract of paper | 1a: Aim | Abstract |
|  | 1b: Methods | Abstract |
|  | 1c: Results | Abstract |
|  | 1d: Conclusions | Abstract |
|  | 1e: Keywords | Keywords |
| Section 2: Background to paper | 2a: Definition | Introduction |
|  | 2b: Theoretical underpinnings | Introduction |
|  | 2c: Concepts and theory development | Introduction |
| Section 3: Aims of paper | 3: Aim | 1 Introduction  2.1.1 Aims |
| Section 4: Methods of paper | 4a: Design | 1.Introduction and Extended Materials 5.1   2.1 Methods /Design |
|  | 4b: People involved | Sections 2.1.3.1; Authors’ Contributions. |
|  | 4c: Stages of involvement | Throughout – “Public and patient / lived experience involvement” section and Introductio; nMethods/Design (1.1 and 2.1) |
|  | 4d: Level or nature of involvement | Preparation of this paper and all components of design have been lived experience led and co-produced. The level and nature of involvement is described throughout the paper, but particular in methods/design (1 and 2.1) and in Author’s Contributions. |
| Section 5: Capture or measurement of PPI impact | 5a: Qualitative evidence of impact | Some informal qualitative evidence was gathered and reported in 2.1.7, with full data supplied in Supplementary Materials “Lived experience perspectives on Impact Log design and dissemination process: additional survey responses” and the protocol contains plans for quantitative data collection. |
|  | 5b: Quantitative evidence of impact | No quantitative evidence gathered during preparation of protocol, but protocol contains plans for quantitative data collection. |
|  | 5c: Robustness of measure | The planned measures have been co-produced, are evidence-based, and are being evaluated during the pilot described in the protocol. |
| Section 6: Economic assessment | 6: Economic assessment | 2.1.3.1 describes the costs associated with this project and there is discussion of the economic implications in the Strengths and Limitations section of the Discussion. |
| Section 7: Study results | 7a: Outcomes of PPI | Given that this is a protocol for a LE-led and co-designed tool, all components involved in Section 7 and 8 are relevant throughout the paper, and more details about individual sections can be found above.  In addition, Section 2.1.7 provides the results and analysis of a survey amongst LE researchers involved in the design of this project and preparation of the manuscript. |
|  | 7b: Impacts of PPI |  |
|  | 7c: Context of PPI |  |
|  | 7d: Process of PPI |  |
|  | 7ei: Theory development |  |
|  | 7eii: Theory development |  |
|  | 7f: Measurement |  |
|  | 7g: Economic assessment |  |
| Section 8: Discussion and conclusions | 8a: Outcomes |  |
|  | 8b: Impacts |  |
|  | 8c: Definition |  |
|  | 8d: Theoretical underpinnings |  |
|  | 8e: Context |  |
|  | 8f: Process |  |
|  | 8g: Measurement and capture of PPI impact |  |
|  | 8h: Economic assessment |  |
|  | 8i: Reflections/critical perspective | Section 2.1.7 provides the results and analysis of a survey amongst LE researchers involved in the design of this project and preparation of the manuscript. The full dataset for these reflections is in Extended Materials Section 7. |

### 5. Narrative Review of Literature Relating to Impact of LEI

#### 5.1 Narrative Review Search and Screening Strategy

The search and screening strategy was designed to provide an overview, rather than systematic and comprehensive evidence synthesis. TG conducted a PubMed search of title and abstracts on 24^th^ July 2025, using terms combining core concepts of LE involvement and co-production. Search terms were: (“lived experience” OR “co-production” OR “service user involvement” OR “participatory” OR “patient and public involvement”) and impact (“impact” OR “evaluation” OR “outcomes”) and mental health (“mental health” [MeSH] OR “psychiatry”). Results were screened for potential relevance and exported into a shared drive in Excel after deduplication. An iterative process of screening for relevance was carried out by EV, LG, HG, and TG during the data extraction phase, with any uncertainties discussed between at least two team members. A grey literature search based on core concepts identified above was conducted by CE using Chat GPT 4.0 in October 2025 and then checked to identify relevant publicly available materials from sources other than peer reviewed journals. References were exported into Excel by HG, re-checked and screened for relevance by HG, JHe, and TG. All LE team members were asked to review results and add papers or resources of potential relevance, and this LE expert review and reference search process continued throughout manuscript preparation, with additional papers integrated into analysis as it progressed.

#### 5.2 Narrative Review Data Extraction and Synthesis

A data extraction table created in Excel by LG was shared with EV and TG for discussion and approval, and adapted for grey literature by HG. This included study date, setting, and participants (for non-review articles); focus and key findings; how impact is defined and measured and main suggestions for increasing impact. Data extraction of peer-reviewed literature was shared between LG and EV, and grey literature between HG and JHe.

The process of narrative synthesis detailed in the main article (1.1) was informal, insofar as a single researcher (TG) created and populated codes and themes, checking and adapting with review team members at various stages.

#### 5.3 Narrative Review Themes Table

| **Themes and Subthemes** | **Articles including themes** |
| --- | --- |
| **How is the impact of Lived Experience Involvement defined and understood?** | |
| **Theme 1: Change to research / service processes and outcomes** |  |
| Increased quality, relevance, and impact of research/intervention | Faithfull, Jennings, Hawke, Papageorgiou, Sheikhan, Riches, Goldsmith, Machin, Jakobsson, Markström, Faulkner, Banfield, Lloyd, Hinterbuchinger, Ghisoni, Hawke 2023, Mjøsund, Sangill, MacInnes, Totzeck, Collins, Musić, McCabe, Hawke 2022, Speyer, Corstens, Faissner, Allen, Sheikhan, Molloy, van der Ham, Springgate, Halvorsrud, Ennis, Veldmeijer, Dray, Hawke 2024, Hawke 2023, Kumpf, McLure, Jones 2021, Aggett, Korteisto, Gatera, Ezaydi, Omeni, Laitila, Veldmeijer, Brett, PiiAF Study Team; Staley 2014, Staley 2015, Hope, Richmond. |
| Systemic change (change in structures, processes, policy relating to research and/or services) | Soklaridis, Siston, Lee, van Draanen; Yoeli, Lloyd, Rose 2018, de Alcântara Mendes, Rose 2014, Åkerblom, Sunkel, Hawke 2023, Rose 2016, Roennfeldt, Colder Carras, Totzeck, Rutherford, Abayneh, Collins, Hawke 2022, Okoroji, Molloy, Trimmel, van der Ham, Springgate, Halvorsrud, Stanyon, Ennis, Veldmeijer, Hawke 2023, McLure, Jones 2021, Loughhead; Alfia-Burstein; Sartor, Lipinski, Rose 2014, Rose 2024, Kortteisto, Faulkner, Banfield, Eyzadi, Laitila, Markström, Staley 2015, |
| Changes to research/intervention design and execution (including design, analysis, outcomes, dissemination, and use). | Faithfull, Jennings, Hawke, Fraser, Papageorgiou, Sheikhan, Goldsmith, Machin, Jakobsson, Markström.Faulkner, Banfield, van Draanen; Hinterbuchinger,Mjøsund, Musić, Okoroji, Allen, Stanyon, Kumpf, Aggett, Brett, PiiAF Study Team, Staley 2015, |
| **Theme 2: Interpersonal and cultural change** |  |
| Change in researcher/clinician attitudes (including reduced stigma and increased understanding, accpetance, and appreciation of mental illness and co-production) | Kortteisto, Colder Carras, Molloy, Riches, Stanyon, Hinterbuchinger, Sheikhan, Trimmel, Hawke 2024, Murphy, Byrne, Staley 2015, Richmond |
| Relationship and epistemic changes within teams (including opportunities for more recognised LE roles and possibilities for LE leadership) | Erikkson, Yoeli, Lipinski, Sunkel, Rose 2024, Haarmans, Hawke 2023, Vescey, Rose 2016, Sangill, Roennfeldt, Byrne (opportunities for meaningful contribution, Colder Carras, Bellingham, Gupta 2023, Musić, McCabe, Hawke 2022, Adams, Faissner, Hawke 2024, Jones 2021, Loughhead, Alfia-Burstein; Aggett, Soklaridis, Fraser, Riches, Goldsmith, Machin, Jakobsson, Siston, Lee, Markström, Stanyon, Rose 2018, Haarmans, Ghisoni, Vesceym , Mjøsund, MacInnes, Roennfeldt, Bellingham, McCabe, Okoroji, Faissner, Allen, Trimmel, van der Ham, Veldmeijer, McLure, Loughhead; Richmond. |
| Increased LE empowerment and opportunities (including all opportunities for personal development amongst people with LE through co-production) | Faithfull, Jennings, Hawke, Papageorgiou, Sheikhan, Siston, Markström, Faulkner, Banfield, van Draanen, Yoeli, Lloyd, Hinterbuchinger, de Alcântara Mendes, Åkerblom, Haarmans, Ghisoni, Hawke 2023, Vescey, Mjøsund, Omeni, Laitila, Gupta 2023, Rutherford, Bellingham, Abayneh, McCabe, Corstens, Okoroji, Faissner, Allen, Sheikhan., Molloy, Trimmel, van der Ham, Springgate, Halvorsrud, Stanyon, Ennis, Veldmeijer, Dray, Hawke 2024, Hawke 2023, Honey, Alfia-Burst; Loughhead; Aggett, McPin LE Group Member, Staley 2015, Richmond. |
| **Methods of impact evaluation** | |
| **Theme 1: Methodologies for assessing impact** |  |
| Qualitative reflections (with an emphasis on gathering data which is primarily reflective and qualitative, rather than quantitative, often without clarity about whether reflections come from LE or non-LE researchers). | Jennings, Hawke, Fraser, Papageorgiou, Riches, Goldsmith, Machin., Jakobsson, Siston, Lee, Faulkner, Stanyon, van Draanen, Yoeli, Lloyd, Hinterbuchinger, Rose 2018, de Alcântara Mendes, Lipinski, Åkerblom, Sunkel, Haarmans, Hawke 2023, Vescey, Mjøsund, Omeni, Rose 2016, Sangill, Laitila, MacInnes, Bellingham, Gupta 2023, Abayneh, McCabe, Faissner, Allen, Sheikhan, Trimmel, van der Ham, Springgate, Halvorsrud, Stanyon, Ennis, Veldmeijer, Dray, Hawke 2024, Hawke 2023, Kumpf, Honey, Alfia-Burst, Loughhead, Alfia-Burstein, Staley 2014, Richmond. |
| Using specific methods for data collection and analysis (including mixed-methods , online surveys, ethographic analysis of Lived Experience Involvement practices) | Ennis, Lee, Erikkson, PiiAF Study Team, |
| Using tools designed specifically to assess co-production. | Totzeck, Rutherford; Collins, PiiAF structure, Honey; |
| Input from non-LE researchers and clinicians | Kortteisto, Faithfull, Molloy |
| **Theme 2: Process indicators** (included measuring impact through tangible changes such as equal pay, increased flexibility, or inclusion of LE leadership). | Soklaridis ,Markström, Banfield, Sartor, Yoeli, Lipinski,Rose 2014, Åkerblom, Sunkel, Haarmans, Hawke 2023, Vescey, Rose 2016, Sangill, Colder Carras, Gupta 2023, Abayneh, Musić, Hawke 2022, Speyer, Corstens, Adams, Faissner, Halvorsrud, Ennis, Hawke 2023, Loughhead, Alfia-Burstein, |
| **Theme 3: The need for alternative ways of understanding impac**t (measuring the impact of Lived Experience Involvement must depart from or critique conventional models of understanding impact , looking for evidence of relational and conceptual change in psychiatry or reduced stigma). | Soklaridis, Laitila, Bellingham, Musić, Speyer, Corstens, Sheikhan, Molloy, Trimmel, Springgate, Hawke 2023, Rose 2024. |
| **Theme 4: Assessing impact of co-production on outcomes and dissemination** | Murphy, Banfield, Sartor, Rose 2018, Sunkel, Yoeli, Hawke 2023, Mjøsund, Ezaydi, Omeni, Collins, Hawke 2022, Speyer, Corstens, Faissner, Halvorsrud, Kumpf., |
| **How to increase impact** | |
| **Theme 1: Changing systems and environment** |  |
| Environmental adaptations (including providing support, power-sharing, building trust, and flexibility in relation to time, structure, and individual needs). | Soklaridis, Jennings, Fraser, Papageorgiou, Sheikhan, Faithfull, Goldsmith, Siston, Lee, Markström, Faulkner, Stanyon, van Draanen; Sartor, Banfield, Hinterbuchinger, Lipinski, Rose 2014, Haarmans, Vescey, Mjøsund, MacInnes, Colder Carras, Totzeck, Bellingham, Musić, McCabe, Hawke 2022, Okoroji, Allen, Sheikhan, Molloy, Trimmel, van der Ham, Springgate, Halvorsrud, Ennis, Veldmeijer, Dray, Hawke 2024, Hawke 2023, McLure, Alfia-Burstein, Frederick, Jennings, MacInnes, Faissner, Richmond |
| Systemic changes (including: ensuring adquate payment, resources, and time; embedding LE involvement into institutional and educational structures; formal acknowledgement of LE research contributions and expertise through e.g. co-authorship, editorial work, principal or co-investigator roles; adaptation of language use; and integration of user-defined outcomes; incorporating opportunities for research involvement within recovery process; ensuring sustainability through creating a 'pipeline' of LE researchers). | Riches, Lee,Sartor, Lloyd, Hinterbuchinger, Åkerblom, Haarmans, Hawke 2023, Sangill, MacInnes., Lee, Musić, McCabe, Okoroji, Faissner, Springgate, Veldmeijer, Hawke 2024, Jones 2021, Alfia-Burstein, Frederick, Sheikhan, Colder Carras, Rutherford, Adam, Molloy, Kumpf, Corstens, Hawke 2023, Honey, Brett. |
| Training (including training for both LE researchers and for non-LE researchers). | Jennings, Fraser, Riches, Jakobsson, Faulkner, Banfield, Lloyd, Hinterbuchinger, Sunkel, Mjøsund, McCabe, Adams, Sheikhan, Hawke 2024, Kumpf, Jones 2021, Alfia-Burstein, Laitila, Byrne, Bellingham, Okoroji, Dray, Loughhead, Soklaridis, Siston, Lee, Yoeli, Laitila, Collins. |
| Reporting and evaluating Lived Experience Involvement (including reflective practices, opportunities for feedback, and standardised modes of reporting and evaluating involvement). | Machin, Jakobsson, Markström, Ezaydi, Omeni, Sangill, Lee, Colder Carras, Totzeck, Rutherford, Collins PiiAF, McCabe, Speyer, Molloy, Halvorsrud, Stanyon, Ennis, Veldmeijer, Hawke 2024, Hawke 2023, Sheikhan, Lloyd, Adams, Trimmel, Papageorgiou, Goldsmith, Yoeli, Rose 2018, Stanyon, Dray, Brett, Staley 2014, |
| **Theme 2: Recognising the value and diversity of lived experience involvement** |  |
| Full LE researcher integration and recognition (highlighting the need to move away from tokenism or stigma within LE involvement and ensure clarity, equality, transparency, recognition, protection, and clear communication surrounding role, expectations, nature and value of LE researchers and of Lived Experience Involvement. | Gatera, Riches, Lee, Banfield, Sartor, Lloyd, Lipinski, Rose 2014, Åkerblom, Sunkel, Haarmans, Hawke 2023, Vescey, Omeni, Rose 2016, Sangill, Colder Carras, Rutherford, Hawke 2022, Adams, Allen, Sheikhan, Trimmel, van der Ham, Springgate, Stanyon, Ennis, Dray, Hawke 2024, Hawke 2023, Kumpf, McLure, Jones 2021, Frederick, Jennings, Siston, van Drannen, Ezaydi, MacInnes, Byrne, Abayneh, Musić, Faissner, Molloy, Halvorsrud, Veldmeijer, Fraser, Papageorgiou, Riches, Machin, Sartor, Hinterbuchinger, Lipinski, MacInnes, Totzeck, Collins, Jakobsson, Richmond |
| Lived Experience Involvement should be included throughout all stages of research/design and delivery | Goldsmith, Riches, Machin, Jakobsson, Markström, Faulkner, Lloyd, Rose 2018, de Alcântara Mendes, Lipinski, Åkerblom (services), Haarmans, Mjøsund, Ezaydi, Omeni, Sangill, Gupta 2023, Hawke 2022, Corstens, Adams, Okoroji, Faissner, Allen, Sheikhan, Trimmel, van der Ham, Springgate, Ennis, Veldmeijer, Dray, Hawke 2024, Hawke 2023, Kumpf, Brett, |
| Ensuring diverse representation within Lived Experience Involvement in terms of demographics and health conditions. | Murphy, Banfield, Sartor, Rose 2018, Sunkel, Yoeli, Hawke 2023, Omeni, Lee, Hawke 2022, Speyer, Okoroji, Faissner, Dray, Hawke 2024, Siston, Markström, Stanyon, Gupta 2023. |
| Safeguarding critical and authentic LE voice (including critical examination of Lived Experience Involvement frameworks, research models, and confronting epistemic injustice | Erikkson, van Draanen;, Rose 2018, Sunkel, Rose 2024, Hawke 2023, Rose 2016; Sangill, Roennfeldt, Gupta 2023, Speyer, Frederick. |

| **Themes and Subthemes** | **Articles including themes** |
| --- | --- |
| **How is the impact of Lived Experience Involvement is defined and understood?** | |
| **Theme 1: Change to research / service processes and outcomes** |  |
| Increased quality, relevance, and impact of research/intervention | Faithfull, Jennings, Hawke, Papageorgiou, Sheikhan, Riches, Goldsmith, Machin, Jakobsson, Markström, Faulkner, Banfield, Lloyd, Hinterbuchinger, Ghisoni, Hawke 2023, Mjøsund, Sangill, MacInnes, Totzeck, Collins, Musić, McCabe, Hawke 2022, Speyer, Corstens, Faissner, Allen, Sheikhan, Molloy, van der Ham, Springgate, Halvorsrud, Ennis, Veldmeijer, Dray, Hawke 2024, Hawke 2023, Kumpf, McLure, Jones 2021, Aggett, Korteisto, Gatera, Ezaydi, Omeni, Laitila, Veldmeijer, Brett, PiiAF Study Team; Staley 2014, Staley 2015, Hope, Richmond. |
| Systemic change (change in structures, processes, policy relating to research and/or services) | Soklaridis, Siston, Lee, van Draanen; Yoeli, Lloyd, Rose 2018, de Alcântara Mendes, Rose 2014, Åkerblom, Sunkel, Hawke 2023, Rose 2016, Roennfeldt, Colder Carras, Totzeck, Rutherford, Abayneh, Collins, Hawke 2022, Okoroji, Molloy, Trimmel, van der Ham, Springgate, Halvorsrud, Stanyon, Ennis, Veldmeijer, Hawke 2023, McLure, Jones 2021, Loughhead; Alfia-Burstein; Sartor, Lipinski, Rose 2014, Rose 2024, Kortteisto, Faulkner, Banfield, Eyzadi, Laitila, Markström, Staley 2015, |
| Changes to research/intervention design and execution (including design, analysis, outcomes, dissemination, and use). | Faithfull, Jennings, Hawke, Fraser, Papageorgiou, Sheikhan, Goldsmith, Machin, Jakobsson, Markström.Faulkner, Banfield, van Draanen; Hinterbuchinger,Mjøsund, Musić, Okoroji, Allen, Stanyon, Kumpf, Aggett, Brett, PiiAF Study Team, Staley 2015, |
| **Theme 2: Interpersonal and cultural change** |  |
| Change in researcher/clinician attitudes (including reduced stigma and increased understanding, accpetance, and appreciation of mental illness and Lived Experience Involvement) | Kortteisto, Colder Carras, Molloy, Riches, Stanyon, Hinterbuchinger, Sheikhan, Trimmel, Hawke 2024, Murphy, Byrne, Staley 2015, Richmond |
| Relationship and epistemic changes within teams (including opportunities for more recognised LE roles and possibilities for LE leadership) | Erikkson, Yoeli, Lipinski, Sunkel, Rose 2024, Haarmans, Hawke 2023, Vescey, Rose 2016, Sangill, Roennfeldt, Byrne (opportunities for meaningful contribution, Colder Carras, Bellingham, Gupta 2023, Musić, McCabe, Hawke 2022, Adams, Faissner, Hawke 2024, Jones 2021, Loughhead, Alfia-Burstein; Aggett, Soklaridis, Fraser, Riches, Goldsmith, Machin, Jakobsson, Siston, Lee, Markström, Stanyon, Rose 2018, Haarmans, Ghisoni, Vesceym , Mjøsund, MacInnes, Roennfeldt, Bellingham, McCabe, Okoroji, Faissner, Allen, Trimmel, van der Ham, Veldmeijer, McLure, Loughhead; Richmond. |
| Increased LE empowerment and opportunities (including all opportunities for personal development amongst people with LE through Lived Experience Involvement) | Faithfull, Jennings, Hawke, Papageorgiou, Sheikhan, Siston, Markström, Faulkner, Banfield, van Draanen, Yoeli, Lloyd, Hinterbuchinger, de Alcântara Mendes, Åkerblom, Haarmans, Ghisoni, Hawke 2023, Vescey, Mjøsund, Omeni, Laitila, Gupta 2023, Rutherford, Bellingham, Abayneh, McCabe, Corstens, Okoroji, Faissner, Allen, Sheikhan., Molloy, Trimmel, van der Ham, Springgate, Halvorsrud, Stanyon, Ennis, Veldmeijer, Dray, Hawke 2024, Hawke 2023, Honey, Alfia-Burst; Loughhead; Aggett, McPin LE Group Member, Staley 2015, Richmond. |
| **Methods of impact evaluation** | |
| **Theme 1: Methodologies for assessing impact** |  |
| Qualitative reflections (with an emphasis on gathering data which is primarily reflective and qualitative, rather than quantitative, often without clarity about whether reflections come from LE or non-LE researchers). | Jennings, Hawke, Fraser, Papageorgiou, Riches, Goldsmith, Machin., Jakobsson, Siston, Lee, Faulkner, Stanyon, van Draanen, Yoeli, Lloyd, Hinterbuchinger, Rose 2018, de Alcântara Mendes, Lipinski, Åkerblom, Sunkel, Haarmans, Hawke 2023, Vescey, Mjøsund, Omeni, Rose 2016, Sangill, Laitila, MacInnes, Bellingham, Gupta 2023, Abayneh, McCabe, Faissner, Allen, Sheikhan, Trimmel, van der Ham, Springgate, Halvorsrud, Stanyon, Ennis, Veldmeijer, Dray, Hawke 2024, Hawke 2023, Kumpf, Honey, Alfia-Burst, Loughhead, Alfia-Burstein, Staley 2014, Richmond. |
| Using specific methods for data collection and analysis (including mixed-methods , online surveys, ethographic analysis of Lived Experience Involvement practices) | Ennis, Lee, Erikkson, PiiAF Study Team, |
| Using tools designed specifically to assess Lived Experience Involvement. | Totzeck, Rutherford; Collins, PiiAF structure, Honey; |
| Input from non-LE researchers and clinicians | Kortteisto, Faithfull, Molloy |
| **Theme 2: Process indicators** (included measuring impact through tangible changes such as equal pay, increased flexibility, or inclusion of LE leadership). | Soklaridis ,Markström, Banfield, Sartor, Yoeli, Lipinski,Rose 2014, Åkerblom, Sunkel, Haarmans, Hawke 2023, Vescey, Rose 2016, Sangill, Colder Carras, Gupta 2023, Abayneh, Musić, Hawke 2022, Speyer, Corstens, Adams, Faissner, Halvorsrud, Ennis, Hawke 2023, Loughhead, Alfia-Burstein, |
| **Theme 3: The need for alternative ways of understanding impac**t (measuring the impact of Lived Experience Involvement must depart from or critique conventional models of understanding impact , looking for evidence of relational and conceptual change in psychiatry or reduced stigma). | Soklaridis, Laitila, Bellingham, Musić, Speyer, Corstens, Sheikhan, Molloy, Trimmel, Springgate, Hawke 2023, Rose 2024. |
| **Theme 4: Assessing impact of Lived Experience Involvement on outcomes and dissemination** | Murphy, Banfield, Sartor, Rose 2018, Sunkel, Yoeli, Hawke 2023, Mjøsund, Ezaydi, Omeni, Collins, Hawke 2022, Speyer, Corstens, Faissner, Halvorsrud, Kumpf., |
| **How to increase impact** | |
| **Theme 1: Changing systems and environment** |  |
| Environmental adaptations (including providing support, power-sharing, building trust, and flexibility in relation to time, structure, and individual needs). | Soklaridis, Jennings, Fraser, Papageorgiou, Sheikhan, Faithfull, Goldsmith, Siston, Lee, Markström, Faulkner, Stanyon, van Draanen; Sartor, Banfield, Hinterbuchinger, Lipinski, Rose 2014, Haarmans, Vescey, Mjøsund, MacInnes, Colder Carras, Totzeck, Bellingham, Musić, McCabe, Hawke 2022, Okoroji, Allen, Sheikhan, Molloy, Trimmel, van der Ham, Springgate, Halvorsrud, Ennis, Veldmeijer, Dray, Hawke 2024, Hawke 2023, McLure, Alfia-Burstein, Frederick, Jennings, MacInnes, Faissner, Richmond |
| Systemic changes (including: ensuring adquate payment, resources, and time; embedding LE involvement into institutional and educational structures; formal acknowledgement of LE research contributions and expertise through e.g. co-authorship, editorial work, principal or co-investigator roles; adaptation of language use; and integration of user-defined outcomes; incorporating opportunities for research involvement within recovery process; ensuring sustainability through creating a 'pipeline' of LE researchers). | Riches, Lee,Sartor, Lloyd, Hinterbuchinger, Åkerblom, Haarmans, Hawke 2023, Sangill, MacInnes., Lee, Musić, McCabe, Okoroji, Faissner, Springgate, Veldmeijer, Hawke 2024, Jones 2021, Alfia-Burstein, Frederick, Sheikhan, Colder Carras, Rutherford, Adam, Molloy, Kumpf, Corstens, Hawke 2023, Honey, Brett. |
| Training (including training for both LE researchers and for non-LE researchers). | Jennings, Fraser, Riches, Jakobsson, Faulkner, Banfield, Lloyd, Hinterbuchinger, Sunkel, Mjøsund, McCabe, Adams, Sheikhan, Hawke 2024, Kumpf, Jones 2021, Alfia-Burstein, Laitila, Byrne, Bellingham, Okoroji, Dray, Loughhead, Soklaridis, Siston, Lee, Yoeli, Laitila, Collins. |
| Reporting and evaluating Lived Experience Involvement (including reflective practices, opportunities for feedback, and standardised modes of reporting and evaluating involvement). | Machin, Jakobsson, Markström, Ezaydi, Omeni, Sangill, Lee, Colder Carras, Totzeck, Rutherford, Collins PiiAF, McCabe, Speyer, Molloy, Halvorsrud, Stanyon, Ennis, Veldmeijer, Hawke 2024, Hawke 2023, Sheikhan, Lloyd, Adams, Trimmel, Papageorgiou, Goldsmith, Yoeli, Rose 2018, Stanyon, Dray, Brett, Staley 2014, |
| **Theme 2: Recognising the value and diversity of lived experience involvement** |  |
| Full LE researcher integration and recognition (highlighting the need to move away from tokenism or stigma within LE involvement and ensure clarity, equality, transparency, recognition, protection, and clear communication surrounding role, expectations, nature and value of LE researchers and of Lived Experience Involvement. | Gatera, Riches, Lee, Banfield, Sartor, Lloyd, Lipinski, Rose 2014, Åkerblom, Sunkel, Haarmans, Hawke 2023, Vescey, Omeni, Rose 2016, Sangill, Colder Carras, Rutherford, Hawke 2022, Adams, Allen, Sheikhan, Trimmel, van der Ham, Springgate, Stanyon, Ennis, Dray, Hawke 2024, Hawke 2023, Kumpf, McLure, Jones 2021, Frederick, Jennings, Siston, van Drannen, Ezaydi, MacInnes, Byrne, Abayneh, Musić, Faissner, Molloy, Halvorsrud, Veldmeijer, Fraser, Papageorgiou, Riches, Machin, Sartor, Hinterbuchinger, Lipinski, MacInnes, Totzeck, Collins, Jakobsson, Richmond |
| Lived Experience Involvement should be included throughout all stages of research/design and delivery | Goldsmith, Riches, Machin, Jakobsson, Markström, Faulkner, Lloyd, Rose 2018, de Alcântara Mendes, Lipinski, Åkerblom (services), Haarmans, Mjøsund, Ezaydi, Omeni, Sangill, Gupta 2023, Hawke 2022, Corstens, Adams, Okoroji, Faissner, Allen, Sheikhan, Trimmel, van der Ham, Springgate, Ennis, Veldmeijer, Dray, Hawke 2024, Hawke 2023, Kumpf, Brett, |
| Ensuring diverse representation within Lived Experience Involvement in terms of demographics and health conditions. | Murphy, Banfield, Sartor, Rose 2018, Sunkel, Yoeli, Hawke 2023, Omeni, Lee, Hawke 2022, Speyer, Okoroji, Faissner, Dray, Hawke 2024, Siston, Markström, Stanyon, Gupta 2023. |
| Safeguarding critical and authentic LE voice (including critical examination of Lived Experience Involvement frameworks, research models, and confronting epistemic injustice | Erikkson, van Draanen;, Rose 2018, Sunkel, Rose 2024, Hawke 2023, Rose 2016; Sangill, Roennfeldt, Gupta 2023, Speyer, Frederick. |

### 6 Origins of the Impact Log: the Mental Health and Justice Project.

The BG Hub *Impact Log* is based on versions designed and used in the multidisciplinary five-year Wellcome Trust-funded Mental Health and Justice project. As dual expertise researchers and Co-Leads of the Service User Advisory Group (SUAG, eight people with LE of mental health conditions), TG and TK created a simple Word document to record how service user input impacted on research (see Supplementary Materials 7). This form was not included in initial project design and protocol.

In *Impact Log* forms circulated to research team members after attending SUAG meetings, researchers were asked to indicate: the questions they asked the SUAG, SUAG input, how SUAG input impacted on their research, with options for additional details, and a final question requesting a brief indication of their general experience. Data was qualitative, non-anonymised, and was not collected from SUAG members. SUAG members were offered opportunities to participate in data analysis and TG, who has expertise in qualitative methodologies, provided basic training in thematic analysis. Initial coding was completed by 6 SUAG members.

Due to time and financial constraints, final analysis and dissemination were not completed. However, this initial version of the *Impact Log* indicated that both LE and non-LE researchers were keen to participate in providing impact data and in data analysis. Initial analysis strongly suggested SUAG consultation had great value, including increased rigour, relevance and feasibility within various research stages, which often exceeded researchers’ initial expectations about potential depth and scope of LE involvement.

#### 6.1 Mental Health and Justice Impact Log Form

**Work Stream / Researcher**

**Date of SUAG input**

**Research question brought to SUAG**

**SUAG input (please be as specific as possible)**

**How did the SUAG contribution impact on your research (please be as specific as possible)?**

**Optional – any other details about research development/outputs relevant to SUAG contribution.**

**Can you give a brief indication of your general experience working with the SUAG?**

### 7 Lived experience perspectives on Impact Log design and dissemination process: additional survey responses

**What have been your experiences being part of the LEAP and impact log design process?**

My experience of being part of the LEAP has been overwhelmingly positive, whereby discussions on study design and strategy have been collaborative and all perspectives felt valued and incorporated. Bringing in lived experience perspectives adds dimensions and ideas to the research that might otherwise be overlooked if researchers worked in isolation. That diversity of perspective not only enriches the process but also makes any tool, or team, or project, stronger and more impactful.

It's been a really interesting process, and I think both the LEAP and the impact log are really important. Combining my lived experience and my experience as a researcher is new to me and has required some reflection to integrate these but I'm proud to be a part of such an important project. I have not come across a LEAP or impact log like this before and I think it's a crucial tool for complex research projects that focus on mental health. The LEAP meetings have been a great opportunity to hear the views and experiences of other researchers with lived experience and I think our voices together are really powerful and add unique insights into the project.

I have learnt so much in this process. I didn't know too much about the landscape of lived experience in mental health research or how its impact is logged currently. I was interested to see all of the published research in this area. I was asked to do a grey literature review and I learnt lots from doing that. I didn't even know what grey literature was before Tania told me. It is a pleasure and a privilege to be working with so many very knowledgeable people. I look forward to seeing everything coming together, all of the valuable input from the different people involved.

Unable to comment as I joined recently

RE impact log design: It's been a really rewarding process. Working alongside the academic team has really helped equalise that power imbalance that is inherent in researcher-LEAP relationships.  
 
RE LEAP: I feel like a valued member of the LEAP. There seems to be a lot of respect in the relationships that have been formed as a direct result of the LEAP - respect between LEAP members and respect with the academics involved too. I've always felt listened to and like my opinions/points of view are warranted and appreciated.

As a researcher in psychiatry who also lives with bipolar, I have found working with the LEAP, both in general, and, specifically on the impact log design process, to be immensely rewarding, empowering, enjoyable, and uplifting.  In my work, I hear so much about the limitations in terms of functioning and career which occur so widely for those experiencing severe mental illness, that it is wonderful to work with a group of researchers, all of whom are living with these conditions, and yet managing to be excellent and dedicated researchers as well. I feel both proud and privileged to be part of this group. Co-production initiatives are so often hampered by tokenism, power imbalances, and failures to be representative.   But the LEAP and impact log publication team are well integrated into the wider study team and making what is recognised to be an immensely important contribution to the work within the project.  It is nice for me, also, to be part of a research group where I am no longer in the minority by virtue of having LE.

My experiences with LEAP, thus far, have been very positive.  
LEAP allows me to bring my lived experience to the table and contribute in a meaningful way. Lived experiences have inherent value which could be useful in informing research directions.  
 
The impact log design process allows LEAP to identify, record & analyze "impact" delivered by LE involvement in research. Seeing this up close & first-hand brings a lot of hope that LE perspectives have real impact on the ground.

It's been very rewarding to be involved in these processes. There is a real push for more lived experience involvement in the design and conduct of mental health research, and I think this is a good thing. It ensures that the profession is alerted to blind spots in not only service provision but also the patient experience, as many mental health projects are geared towards clinical applications and outcomes.

It has been lovely to get to know other people more and find out lots about loved experience in research.

**What value you think the impact log evaluation tool could add to either the Brain and Genomics Hub project or the mental health research field in general?**

The impact log evaluation tool is especially valuable because it helps to quantify the impact of lived experience advisory work and monitor its influence. This kind of evaluation is really needed in LEAP advisory work, because it allows us to move beyond tokenistic involvement of those with lived experience and identify where lived experience input has the biggest impact, and where it could be strengthened. In turn, that makes the process more transparent, accountable, and ultimately more meaningful—both for researchers and for the communities the research is meant to serve.

While other research projects I have worked on include the voices of people with lived experience, it is often hard to evaluate the impact they have. From my experience as a researcher and as a lived experience member of research projects I've found this especially difficult when the project is complex, like the study being done by the Brain and Genomics hub.  Being able to measure and demonstrate this impact is a really important part of co-designing research and is a important part of the process of participating in research as someone with lived experience.  Developing an impact log evaluation tool is a novel idea that has the chance to do this within the Brain and Genomics Hub and in the mental health research field in general. I think it's so important that people with lived experience are involved in the development of this tool and it's fantastic to be a part of this.

It would be an excellent means of recording the impact of lived experience input into the Brain and Genomics Hub and could be used more widely by other research projects. There doesn't appear to be a tool that is currently consistently used so perhaps this could become that tool, a gold standard for use in mental health research.

Unable to comment as I joined recently

I think the impact log evaluation tool is going to be beneficial in helping us operationalise something that is inherently subjective and often difficult to convey.

I am hopeful that the Impact log evaluation tool could have immense value both for the Hub and for mental health research in general.  Although the field of co-production and lived experience involvement has progressed hugely in recent years, there is still a lack of established ways to measure and demonstrate impact and to evaluate how well lived experience involvement is going within a project. I am hopeful that the IL tool can provide a tool-kit and framework which others will be able to adapt and use within their research, so that we move forward in making co-production a more methodologically developed component of research, as well as being a tool which recognises the interpersonal components of co-production work and the difference this can make to everyone involved.

The impact log evaluation tool will help measure the "impact" of LE involvement in research. This is relevant for all stakeholders of brain & genomics hub project - including funding bodies. In a larger context, the impact log evaluation tool can be utilized across the mental health research field.

The tool constitutes a valuable reference resource for other researchers, as well as the Brain and Genomics Hub itself.

I think it could measure and demonstrate the impact of involving people with lived experience and be used for other projects too for comparison.
